# Minimal algorithms for knowledge representation in clinical decision support systems research: a theoretical and empirical analysis

**DOI:** 10.1101/2022.04.20.22274099

**Authors:** Anderson A. Eduardo, Rafael M. Loureiro, Adriano Tachibana, Pedro V. Netto, Tatiana F. de Almeida, Luiz Henrique Alves Monteiro, André P. dos Santos

## Abstract

Clinical decision support systems (CDSS) figures out as one of the most promising technologies for data-centered and AI-prompted healthcare. Its current developments are mainly guided by two disparate mindsets, namely a machine learning-centered framework and a classical rule-based framework. These respective approaches presents contrastive pros and cons. In the present study we provide an analysis showing that these two mindsets are actually related to each other, and straightforward algorithms are feasible by combining current standards for machine learning and classic decision tables algorithms. A theoretical analysis are provided, as well a computational implementation (in python). A real case scenario on radiological immaging exam prescription is used to ilustrate the successfully application of our results. Future work on benchmarking the proposed algorithms embodied in a fully operational clinical decision support system could extend our findings towards daily used systems.

## 1 Introduction

In the last decade, modern digital technology become pervasive in healthcare and medical systems. In terms of intellectual historical evolution, the ongoing disruption by the emergence of the so called fourth scientific paradigm reverbered in advancing health sciences, existing a long and apparent road of technological advances ahead [6, 8].

In this context, clinical decision support systems (CDSS) have transvased academic circles to daily life, with large operational systems affecting thousand of patients and professionals per day [7]. Today, CDSS is perceived by many authors as one of the most promising concept among emergent technologies in healthcare [5, 2, 7, 1].

In practical terms, CDSS are implemented as a software, generally integrated as a service in previously existing local systems [7]. The software is kept runing under the rotinetely use of a local system. But, internally, prescription of medical exams are processed by the CDSS and some type of signalization is shown to the user, sugesting the best options given patient information [7].

There are different and specific modes of implementation and operationalization of CDSS [4, 7]. Despite of that, they are built upon a same system architecture, comprising [7]: (i) an interface layer, where the user inputs patient data, receive prescription suggestions and queries are parsed; and (ii) the knowledge base, being an algorithm capable of modeling expert knowledge regarding a specific problem or intellectual domain.

The literature on the knowledge base component of CDSS discusses a variety of algorithms, from explicit IF-ELSE rules to deep learning. Naturally, every approach to knowledge modeling will have its pros and cons. But, in our interpretation, research advances have largelly been internally biased for each one of the respective lines of approach, with loosely interaction for theoretical synthesis and integrative developments. Interpretability of knowledge representation is a relevant example, with, on one hand, classic IF-ELSE approaches having clearer representation of internal extructure (directaly affecting the interpretability of systems suggestions), but at a high cost of implementation and maintainance, and, on the other hand, machine learning approaches, having low (or very low) internal interpretability, but with a lesser cost of implementation and maintainence and, importantly, with inherent uncertainity in outputs [7].

Thus, in present study we provide an analysis showing that straightforward algorithms are feasible, combining current standards in massive dataset formation for machine learning and classic decision tables algorithms. We show that there are place for mathematical theory bridging classic and modern approaches for knowledge representation in CDSS and we ilustrate it providing “white-box”, graph-based minimal algorithms, with experimental results for real radiological imaging data.

## 2 The decision table formalism

Strictly speacking, a decision table is a table representing the exhaustive set of mutual exclusive conditional expressions, within a predefined problem area (see [9]).

There are two fundamental sets for a decision table, 𝒞 and 𝒜, namely the *condition set* and the *action set* [9]. The condition set is defined as follows (definition (2.1)).

### Definition 2.1.

The Condition set, 𝒞, consists of *n* elements *c*_*i*_, 1 ≤ *i* ≤ *n*, which are, in turn, formed by the tuple (*c*^⟨*S*⟩^, *C*^⟨*T*⟩^)_*i*_, with 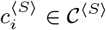 and 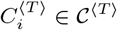, such that

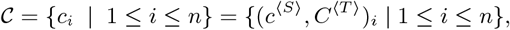

where 𝒞 ^⟨*S*⟩^ are the condition subjects, with each *i*^*th*^ subject having a domain 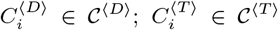 is the set of possible logical states for the *i*^*th*^ subject, being stated in terms of 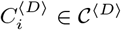 and mapping to a one and only one item of the cartesian product 𝒜^⟨*V*⟩^ × 𝒜^⟨*V*⟩^ (which is definied bellow).

For a set 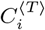, its condition states follows the definition below (definition (2.2)).

### Definition 2.2.

Each set of condition state, 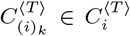, defines a subset 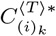, such that 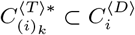 and

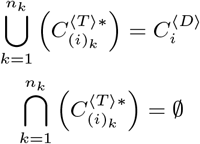

Regarding the action set, it is defined as follows (definition (2.3)).

### Definition 2.3.

Similarlly to the Condition set, the Action set, 𝒜, is formed of *m* actions *a*_*j*_, 1 ≤ *j* ≤ *m*, being *a*_*j*_ = (*a*^⟨*S*⟩^, *A*^⟨*V*⟩^)_*j*_, with 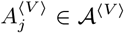 the set of possible values for the *j*^*th*^ action of 𝒜, and 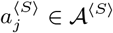 the *j*^*th*^ action item, such that

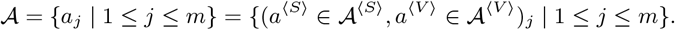

In order to illustrate such concepts in the context of the present paper, we provide the following example.

**Example**. *Consider a decision table in a simplifyied context of medical immage prescription. The Condition set, in terms of* 𝒞^⟨*S*⟩^ = {*age, clinical indication*}, 𝒞^⟨*D*⟩^ = {{0, 1, 2, …}, {*Indication* 1, *Indication* 2}}, 𝒞^⟨*T*⟩^ = {{*age* ≤ 17 → *children, age >* 17 ∧ *age* ≤ 50 → *adult, age >* 50 → *senior*}, {*Indication* 1, *Indication* 2}}, *and* 𝒞^⟨*T* ⟩*^ = {{{1, 2, …, 17}, {18, 19, …, 50}, {51, 52, …}}, {{*Indication* 1}, {*Indication* 2}}} *is defined as*

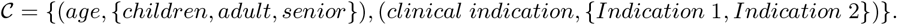

*The Action set, in terms of* 𝒜^⟨*S*⟩^ = {*clinical exam*} *and* 𝒜^⟨*V*⟩^ = {*exam* 1, *exam* 2, *exam* 3} *is defined as*

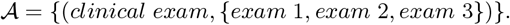

Given a decision table for which these definitions completely holds, we got a decision table function, as stated by the following theorem (theorem (2.1)).

### Theorem 2.1.

*(Decision table function) Considering the cartesian products* 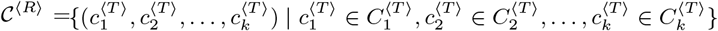 *and* 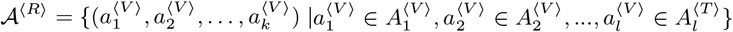, *there is such a function*

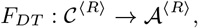

*mapping each action in* 𝒜^⟨*R*⟩^ *from one or more conditions in* 𝒞^⟨*R*⟩^.

*Proof*. Let 𝒳 = {𝒳 | *x* ∈ 𝒞^⟨*R*⟩^} and 𝒴 = {*y* | *y* ∈ 𝒜^⟨*R*⟩^}. Consider a relation *R* ⊂ {(*x, y*) | *x* ∈ 𝒳 *y* ∈ 𝒴}. As 𝒞^⟨*R*⟩^ is defined in terms of 𝒞^⟨*T*⟩^, the definitions (2.1) and (2.2) holds for any element or subset of 𝒞^⟨*R*⟩^. Thus

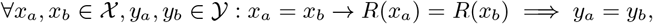

meaning that *R* is a function with domain 𝒳 and image in 𝒴. Consequently, 𝒳 ⊂ 𝒞^⟨*R*⟩^ and 𝒴 ⊂ 𝒜^⟨*R*⟩^ implies that *F*^*DT*^ *is a function*.

Traditionally, a decision table is represented in a (row-driven) augmented matrix, as follows (equations (1), (2) and

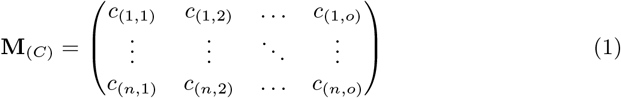

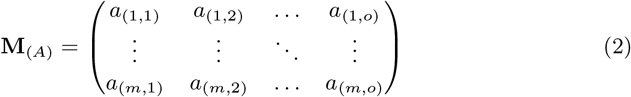

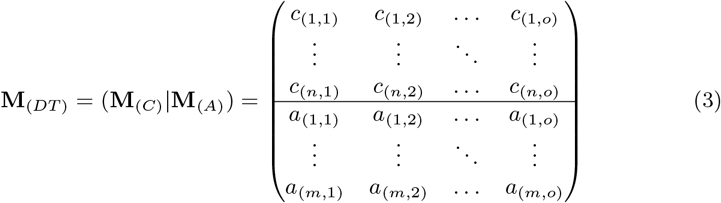

Here, *c*_(*i,l*)_ ∈ 𝒞^⟨*T*⟩^, with *i* ∈ {1, 2, …, *n*} and *l* ∈ {1, 2, …, *o*} refer to condition state values for the *i*^*th*^ condition subject and *l*^*th*^ rule. The values *a*_(*j,l*)_ ∈ 𝒜^⟨*V*⟩^ with *j* ∈ {1, 2, …, *m*} and *l* ∈ {1, 2, …, *o*}, refer to actions for the *j*^*th*^ action subject and *l*^*th*^ rule.

## 3 Linking decision tables and statistical machine learning datasets

Despite of both decision tables and machine learning be cirscunsribed whithin the wide field of artificial intelligence, these subjects has been, conceptually and temporally, relatively far from each other. The decision table formalism were developed in the age of symbolic approaches to artificial intelligence modelling. Machine learning, on the other hand, emerged as a broad theoretical mindset, which became in vogue in the recent decades, becaming the dominant paradigm for the current generation of artificial intelligence researchers and practioners. Despite of such historical bias, mathematical formalisms in the machine learning field and the decision table formalism are not disparate from each other, having place for theoretical synthesis and synergistic developments.

By the current formalism of statistical learning theory for supervised learning, a machine learning problem can be definied as follows (definition 3.1) (see [3]).

### Definition 3.1.

A machine learning problem consists of finding a function capable of mapping a random variable ***Y*** ∈ ℝ from a vector of ***x*** ∈ ***X***, with ***X*** ⊂ ℝ^*n*^. Such a function, defined here as *h*(*x*) : ***X*** → ***Y***, belongs to a hipothetical space of functions, ℋ, and can be computed by the means of minimizing a loss function *L*(*h*(·), *S*) with respect to a parameterized hypothesis, *h*, and considering *S*, a collection of observed instances of ***X*** and ***Y***.

By definition, and as stated by the definition (3.1), a machine learning problem explicitly depends on a collection of empirical observations. Thus, we state a more specific definition for *S*, as follows (definition 3.2).

### Definition 3.2.

Let *Z* be the catesian product ***X*** ×***Y***·A collection of empirical observations, *S*, is a statistical sample over the unknown probability distribution of *Z*, being constituted by tuples (***x*** ^⟨*observed*⟩^, *y*^⟨*observed*⟩^)_*i*_, 1 ≤ *i* ≤ *o*, with the vector 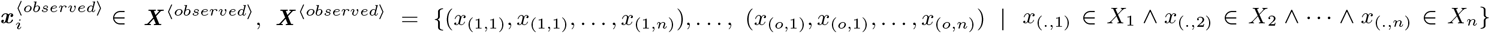, observing that ∀*X*_*i*_ ⊂ ***X*** : ∪_*i*_(*X*_*i*_) = ***X*** ^⟨*observed*⟩^; and with 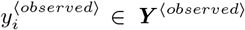 · *S* is parsed by a learning algorithm which implements *L*(*h*(·), *S*) and is capable of returning *h*(·)_*S*_, which refers to a parameterized function regarding a particular sample dataset, *S*. Thus, the parameterization found by the algorithm is fundamentally determined by the sample dataset.

In terms of tabular datasets, ***X*** ^⟨*observed*⟩^ and ***Y*** ^⟨*observed*⟩^ can be represented in a matricial form, as shown below (equations (4) and (5)).

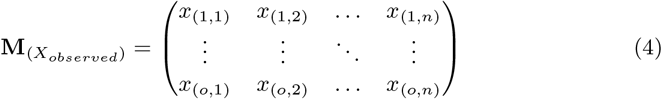

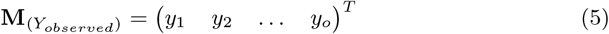

Thus, *S* can be represented by the augmented matrix given below (equation (6)).

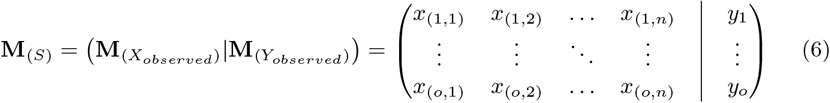

The matricial representation of decision tables is structuturally equivalent to such matricial representation of *S*, **M**_*S*_, as stated by the theorem (3.1).

### Theorem 3.1.

*(The equivalency theorem for* **M**_(*DT*)_ *and* **M**_(*S*)_*) A matricial representation of a decision table*, **M**_(*DT*)_, *given by equation (1), is equivalent to a matricial representation of a statistical machine learning dataset*, **M**_(*S*)_, *given by equation (4), if* **M**_(*DT*)_ *is transposed, such as*

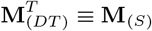

*Proof*. Considering definitions (2.1), (2.2) and (2.3), each one of the *n* subjects in 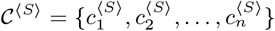 have a domain 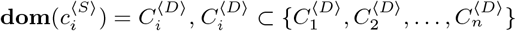, being assigned in a row-driven fashion in the matrix **M**_*C*_, given by equations (1) and (3). Each column *l* in **M**_*C*_ can be conveniently represented as a *n*-size tuple ***t*** _*l*_ composed by state values *v*_*i*_, 1 ≤ *i* ≤ *n*, each one row-wise assigned to its respective subjects, 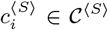. Clearly, the *o*-columns in **M**_*C*_ is relative to the number of decision rules embeded in a decision table and, in turn, informing the number of such tuples, ***t*** _*l*_. Now, consider that each row in **M**_*S*_ groups the values of the *n* features (or traits, or variables, or dimensions) for each empirical observation,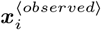. Necessarily, each feature, *X*_*i*_, have a proper domain, **dom**(*X*_*i*_) ⊂ ℝ, and each vector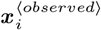, being an empirical observation, by definition, can also be represented as a tuple ***t*** _*l*_ of observed state values *v*_*i*_, column-wise assigned to its respective feature *X*_*i*_, and showing that condition subjects are equivalent of features, or

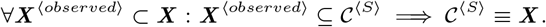

Thus, any hypothetical subject, *C*_*a*_, with its own domain **dom**(*C*_*a*_), can have its state value imputed in a tuple, *t*_*C*_, comprised of *n* state values, along other *n* − 1 state values for other subjects, such as *C*_*b*_, *C*_*c*_, *C*_*d*_, …, *C*_*z*_. An arbitrary number of such tuples can be stacked either in a row-oriented fashion, forming a matrix **M**_1_, or in a column-oriented fashion, forming a matrix **M**_2_, without any loss of generality. It can be easilly seen that

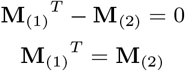

Naturally,

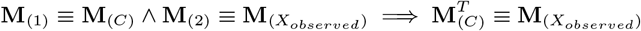

The application of this same rationale for equations (2) and (5) is trivial, proving the theorem (3.1). □

Thus, any dataset as typically arranged in current machine learning practioning is ready to be used for a decision table implementation, as long as it is in accordance with definitions (2.1), (2.2 and (2.3)).

An interesting consequence of theorem (3.1) is stated by the following corollary.

### Corollary 3.1.

*Given a hypothesis h* ∈ ℋ, *any machine learning algorithm, in terms of the definitions (3.1) and (3.2), can learn and furnish a computational representation of any particular decision table function, as stated at theorem (2.1), provided that the loss function tends to zero. This is expressed by*

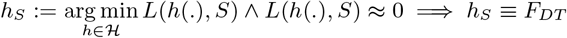

*Proof*. Let a dataset *D* be definied in terms of the equations (4), (5) and (6). Let a function *g*_*Error*_ : ℝ^*n*^ → ℝ be an error function, i.e., a function able to compute an arbitrary degree of correctedness for the output of functions *f*_*i*_ ∈ ℱ, being ℱ a set of particular functions. Now, consider a function *f*_*DT*_ ∈ ℱ, which is specified in terms of the definitions (2.1), (2.2) and (2.3). Also, consider a function *f*_*ML*_ ∈ ℱ, specified in terms of definitions (3.1) and (3.2). By the definitions, we have *g*_*Error*_ (*f*_*DT*_, *D*) = 0, i.e., when applying *f*_*DT*_ to the dataset *D*. Also by the definitions, we have that, for any situations in which *L*(*f*_*ML*_, *D*) ≈ 0 is achievable, we have

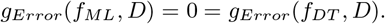

In this cases,

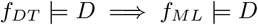

This result makes *f*_*DT*_ and *f*_*ML*_ equivalent in the representation of *D*, or

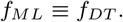

As *f*_*DT*_ is defined is the same terms of *F*_*DT*_ and *f*_*ML*_ is defined in the same terms of *h* ∈ ℋ, we got

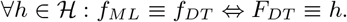

Corollary (3.1) pave the way for applying current machine learning algorithms in computational representation of decision tables in a straightforward fashion. This shows that, as we argued before, these are not rival formalisms, existing potential for theoretical and practical synthesis. Despite of corollary (3.1), we would like to remark that representing a decision table in terms of *h* ∈ ℋ (see definition (3.1)) impels loss of interpretability of internal decision structure, being contingent to algorithm features.

## 4 Flexible decision tables

In the previous section we have shown that decision tables have a good fit with machine learning datasets, making them adherent to the canonical structure for modern datasets, widely used by machine learning practioners. Such result may help to bring it closer to the most palatable algorithms routinely used for artificial intelligence modelling.

However, as stated in the furnished definitions, only one output decision should be expected for any inputted query in a decision table model. Unfortunatelly, it can be too restrictive for many pratical applications. As an example, this is the case for prescription of radiology image exams. Profissionals in this field commonly ends up with more than only one well appropriate radiological immaging options for a same patient condition. So, in order to make simpler algorithms really usefull and adequate in suporting such cases, we should flexibilize decision table formalism.

Here we propose that a more flexible or generalizable background for decision tables can be obtained through a minor changing in the basal definitions at decision table formalism, as stated in the following definition (definition 4.1).

### Definition 4.1.

A decision table can be defined as a ordered triple (𝒞^⟨*R*⟩^, 𝒜^⟨*R*⟩^, *G*(*V, E*)), with 𝒞^⟨*R*⟩^ = 𝒞^⟨*T*⟩^ × 𝒞^⟨*T*⟩^, 𝒜^⟨*R*⟩^ = 𝒜^⟨*V*⟩^ × 𝒜^⟨*V*⟩^ and *G*(*V, E*) being a directed graph, with vertices *V* ⊆ 𝒞^⟨*R*⟩^ ∪ 𝒜^⟨*R*⟩^ and edges *E* ⊆ 𝒞^⟨*R*⟩^ × 𝒜^⟨*R*⟩^, mapping from 𝒞^⟨*R*⟩^ to 𝒜^⟨*R*⟩^. In such terms, a decision table consists in the binary relation, *R*_*DT*_, over sets 𝒞^⟨*R*⟩^ and 𝒜^⟨*R*⟩^, being defined as

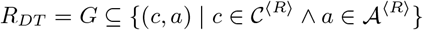

### Theorem 4.1.

*(Flexible decision table) Given definition (4.1), F*_*DT*_ *is a special case of R*_*DT*_.

*Proof*. The proof naturally emerge from the fact that *F*_*DT*_ is a function and *R*_*DT*_ is a mathematical relation. So,

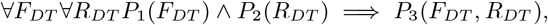

being *P*_1_, *P*_2_ and *P*_3_ the predicate symbol for *is a mathematical function, is a mathematical relation* and *is a special case*, respectivelly. □

With the definition (4.1), we are generalizing definition (2.1) and, consequently, we provide that *F*_*DT*_ (see theorem (2.1)) naturally be a particular case of *R*_*DT*_ (definition (4.1) and theorem (4.1)). This is a subtle change from the classic definition of decision tables, but with consequences making them more interesting for modern practical use, such as decision support systems for radiology exam prescription.

Also, it is important to note that, in the terms of definition (4.1) and theorem (4.1), any results obtained considering decision tables strictly as a mathematical function do not apply to *R*_*DT*_. Here, the corollary (3.1) is the most relevant example.

Fortunately, in modeling a decision table as a mathematical relation and representing it as a graph object, we circunscribe our focal problem in the sound grounds of graph theory. This is very convenient, because algorithms and computational tools specific for such abstract objects are well developed and “white box” models are feasible. In the next sextion, we explore the simpler algorithms for computing *R*_*DT*_.

## 5 Algorithms

In this section, we provide algorithms for the decision table graphs, in accordance to definition (4.1). It is assumed that the input dataset is structured in accordance to definitions (3.1), (3.2) and (4.1), as well as the theorem (3.1).

Given the nature of the dataset for a flexible decision table, it is very convenient to entirely load each vector of condition states and action values (i.e., the rows of **M**_(*S*)_), representing it as a single computational object (e.g., a *n*-tuple). By our definition of flexible decision table, such vectors constitute the paths of a graph *G* for the relation *R*_*DT*_, thus the proper paths of the decision table.

The algorithm (1) shows that paths can be readly obtained from data arranged in terms of equation (6), providing a straightforward computational representation of *G* and, by definition, the relation *R*_*DT*_ for a given decision table.

### Algorithm 1 The *Load data as graph paths* algorithm

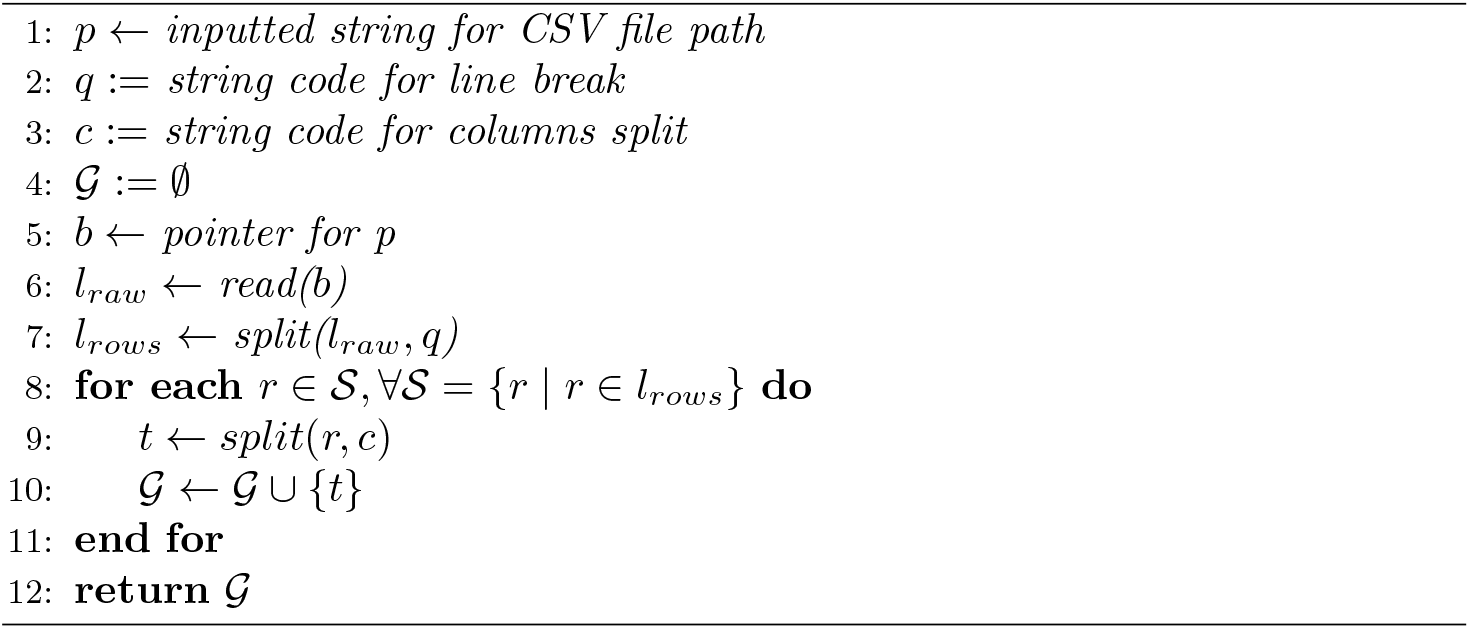

In the context of algorithm (1), *read* is a method for obtaining data in computer physical memmory through a pointer object, and *split* is a method to split a string at specific positions (given by *c*). These methods are commonly provided on most of currently used programming languages for artificial intelligence implementation (e.g., python).

Following, given a graph *G*, represented acconding to algorithm (1), querying for actions, *a*^⟨*V*⟩^ ∈ 𝒜^⟨*V*⟩^, given a particular set of condition states, 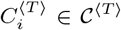, should be possible. In algorithm (2), we show how a set of paths to actions, constrained to inputted condition states, can be efficiently maped.

### Algorithm 2 The *Find paths* algorithm

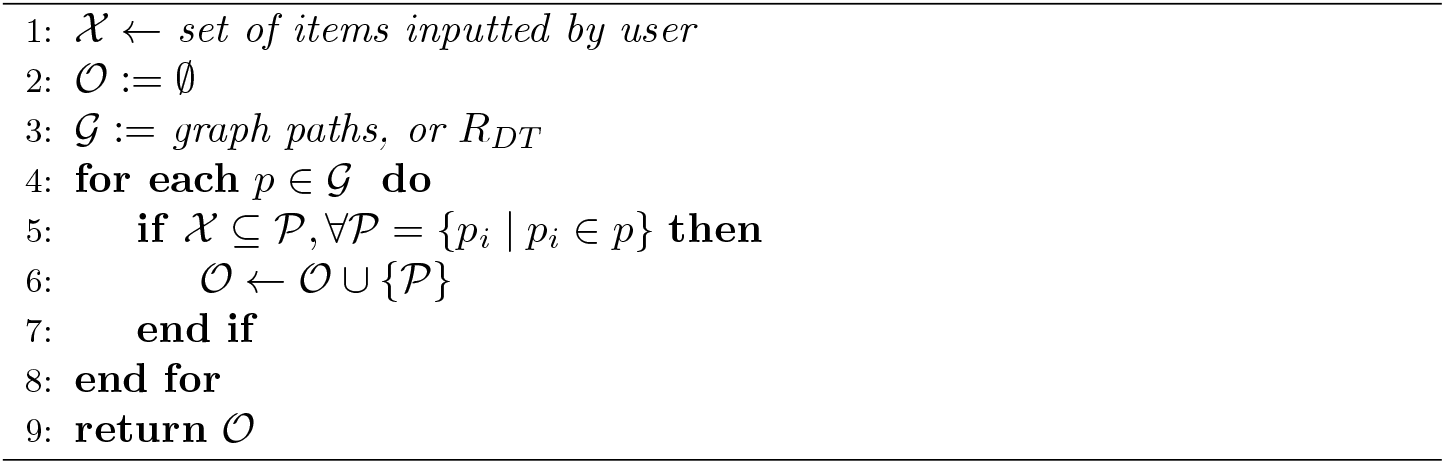

Finally, our representation of *G*, in order to achieve a straightforward representation of *R*_*DT*_, can easilly be rearranged into the classical representation for graph objects. This is especially relevant for the treatment of decision tables within the scope of long standing computational tools available for mathematical operations on graphs. Thus, in algorithm (3) we show that an edge list is readily obtainable for *G*.

### Algorithm 3 The *From paths to edges* algorithm

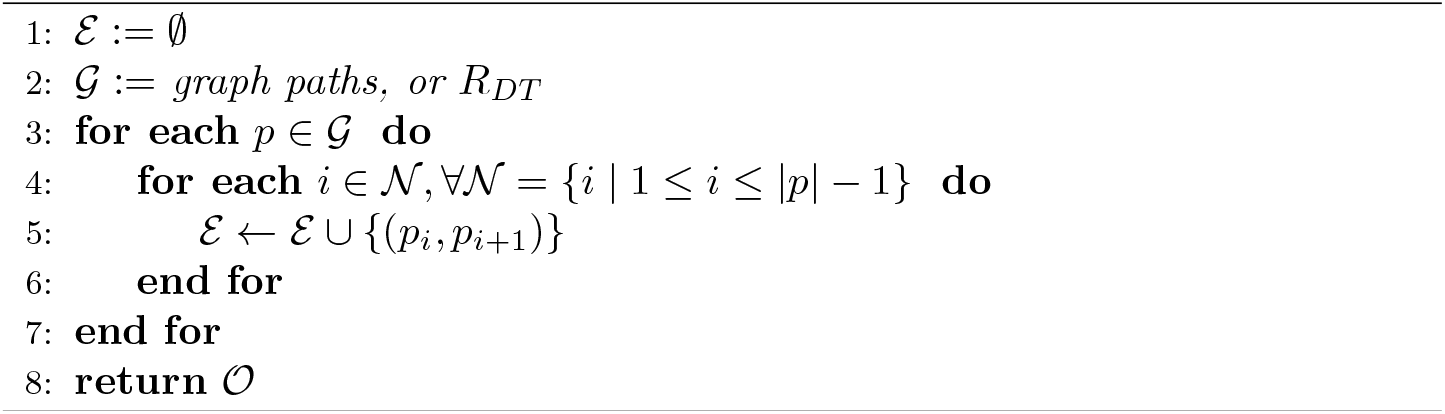

It is easy to see that the temporal complexity of both algorithm (1) and algorithm (2) is *O*(*n*). For algorithm (3), temporal complexity is *O*(*n* × *m*), being 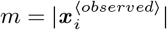.

## 6 Experiments

In order to subdue our theoretical arguments to the scrutiny of reality, in this section we carry out an experiment exploring an application for a clinical decision support system. Focusing on a medical radiological context, we compiled a decision support dataset for immaging exams for head and neck situations (see `Supplementary Material`). Our data were obtained from *Appropriateness Criteria* of American College of radiology (ACR), available online at https://acsearch.acr.org/list.

Thealgorithms (1), (2) and (3) were implemented in pure `python` programming language (version 3.9). Complementarily, we used the package `networkx` (version 2.6.3) to explore the output of algorithm (3).

Bellow, python coding for the mentioned algorithms are shown.

**Listing 1:**
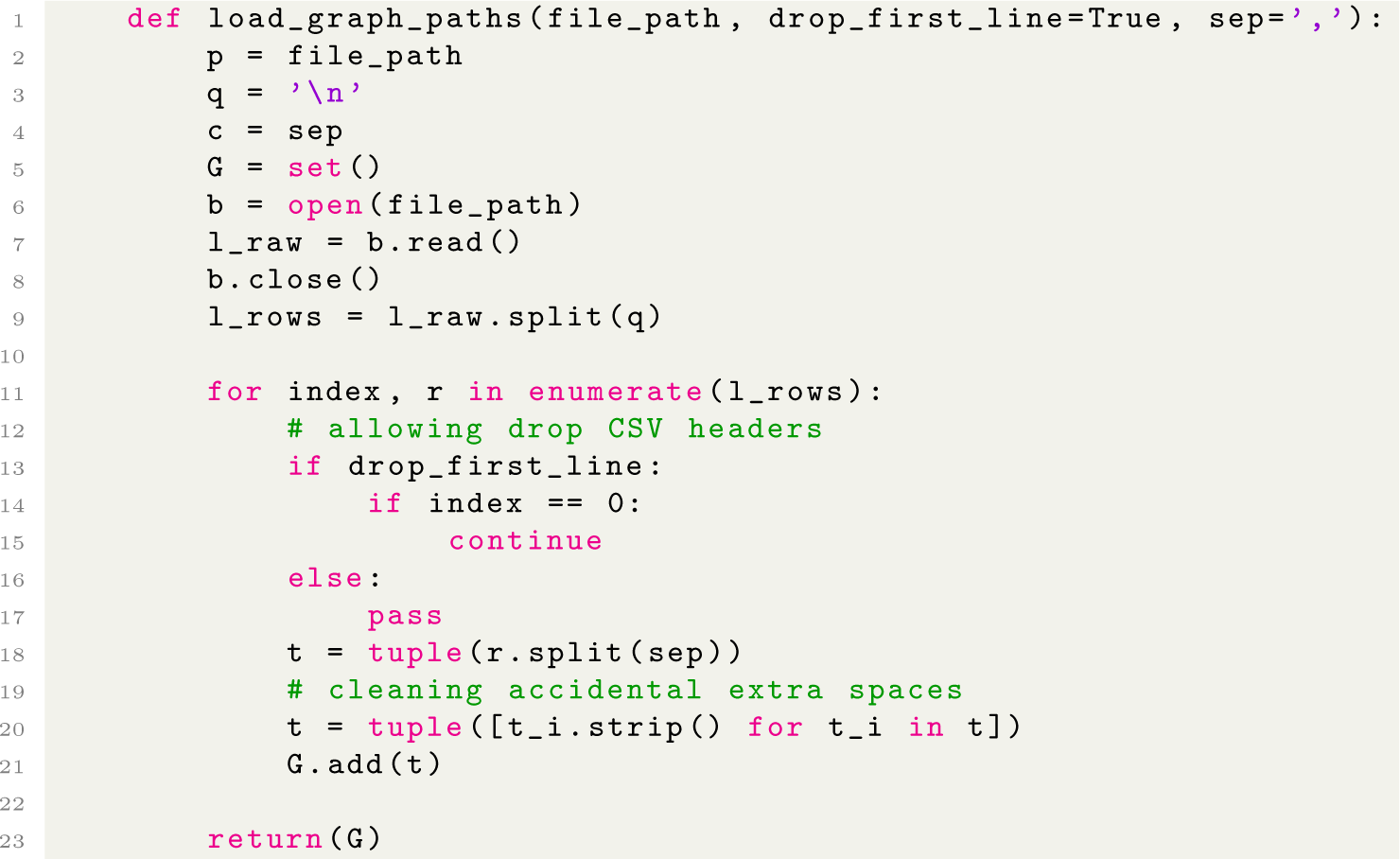
Python implementation of algorithm (1).

**Listing 2:**
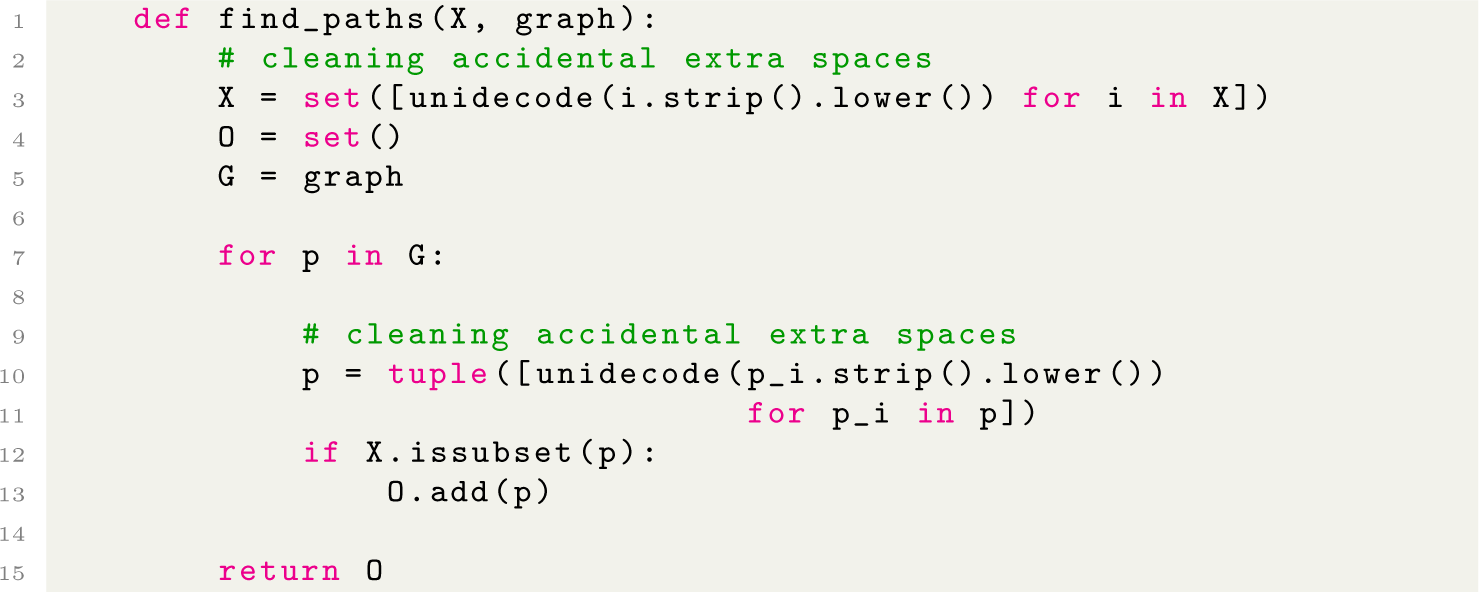
Python implementation of algorithm (2).

**Listing 3:**
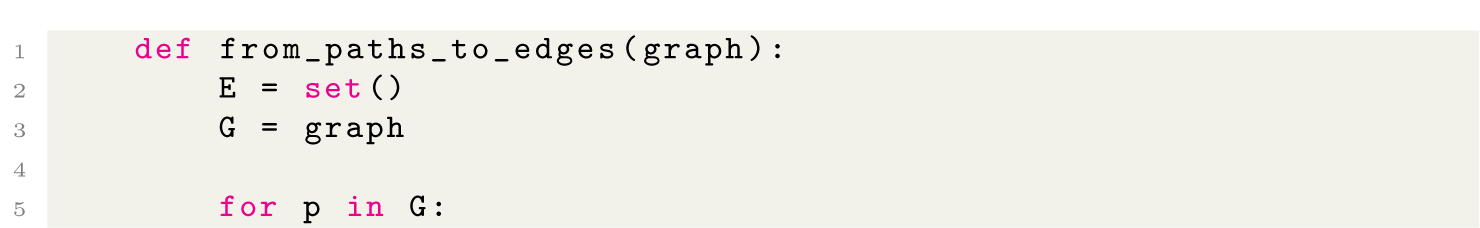
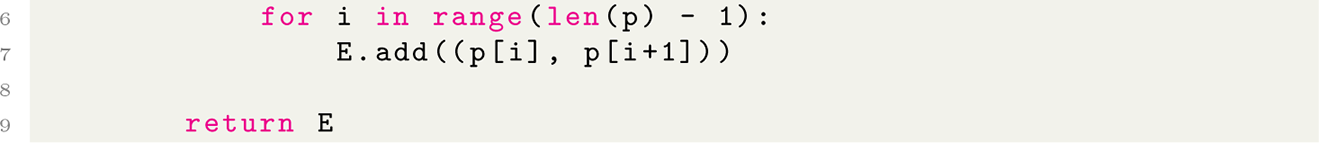
Python implementation of algorithm (3).

The complete implementation of our experiments is provided in Supplementary Material. Bellow, we provide the main results.

## Experiment 1: *obtaining a representation of G for the experimental dataset*

Using the algorithm (1), implemented as the method `load_graph_paths`, the obtained output is shown in listing (4).

**Listing 4:**
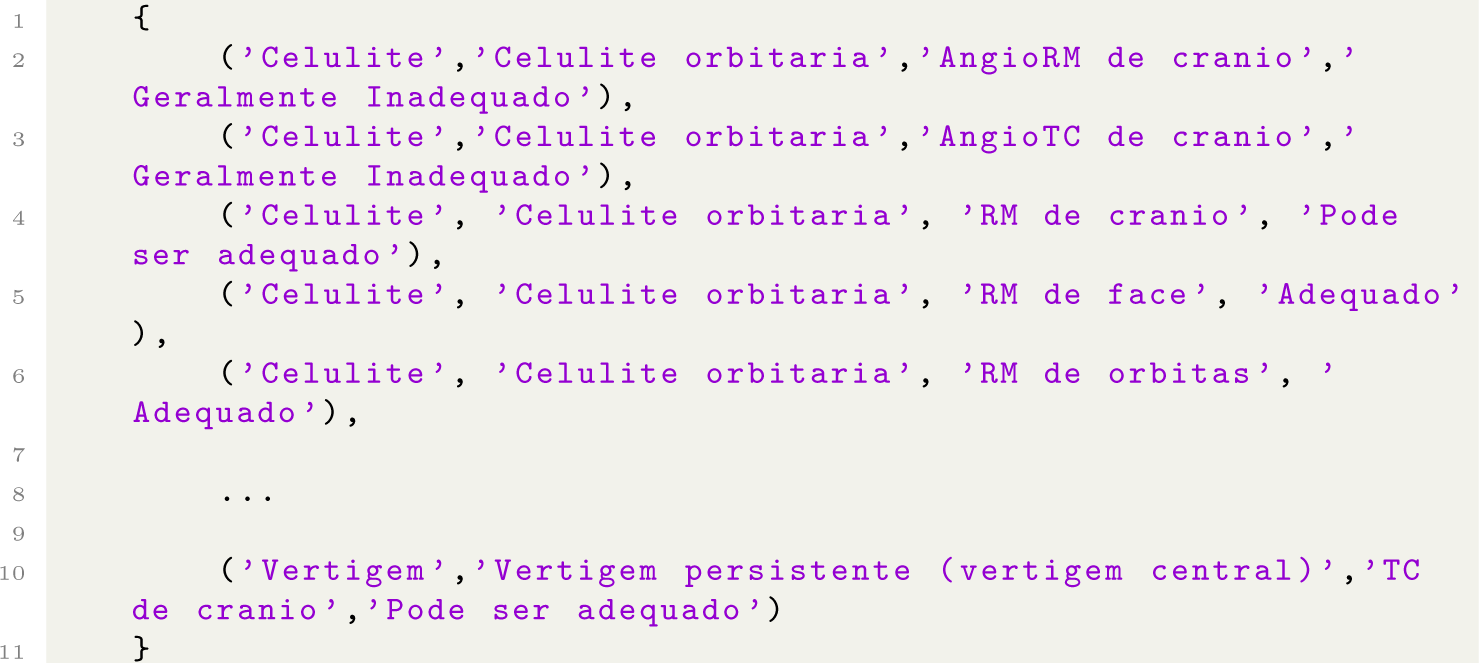
Output for Experiment 1.

## Experiment 2: *maping paths to actions, given input constrains*

Now, with the implementation `find_paths` for algorithm (2), the obtained output for the query `find_paths([‘Vertigem’, ‘Vertigem episodica(vertigem periferica)’])` is shown in listing (5).

**Listing 5:**
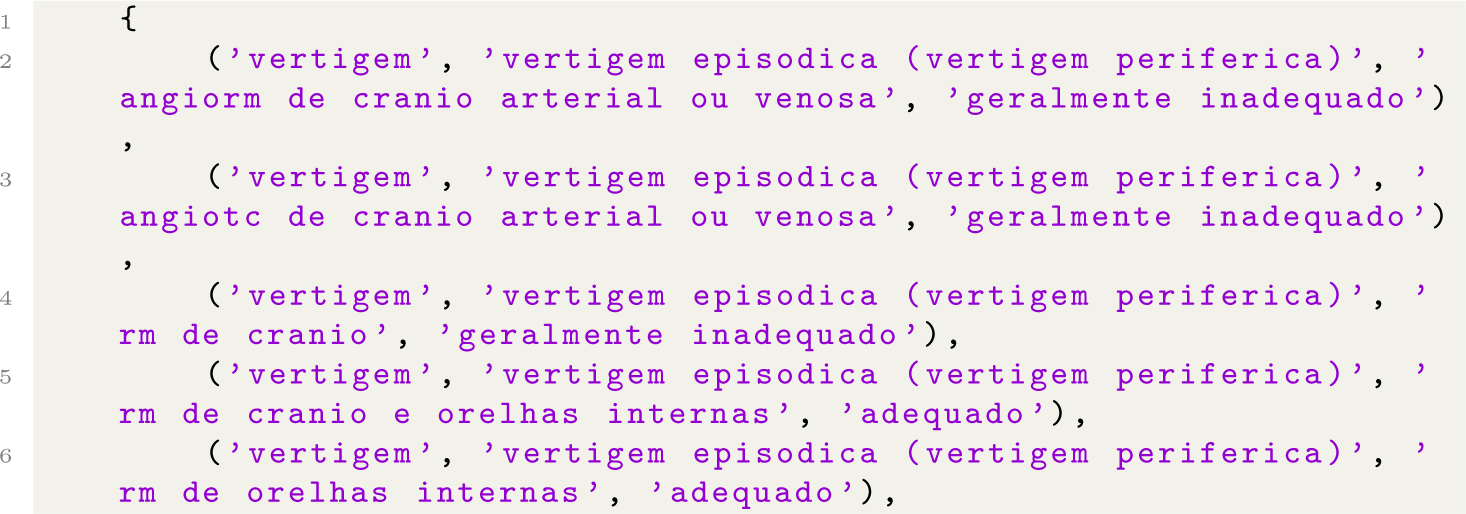
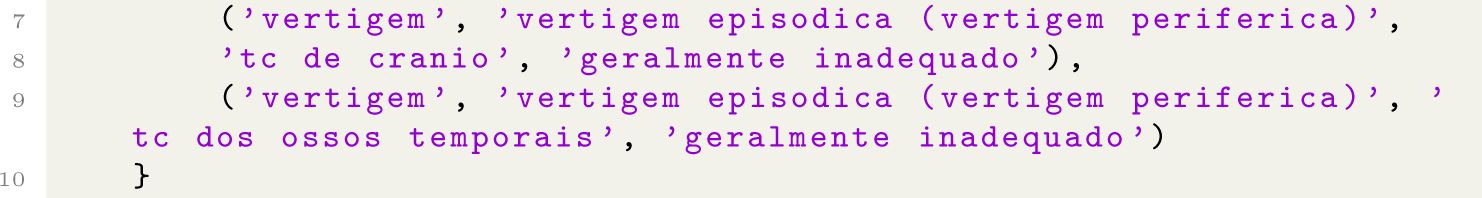
Output obtained for Experiment 2.

**Experiment 3: *obtaining canonical representation of graphs***

With the implementation `from_paths_to_edges` for algorithm (3), the obtained output is depicted on listing (6). This output were succefully parsed by `networkx` methods, being a renderization of *G* provided in figure (6).

**Listing 6:**
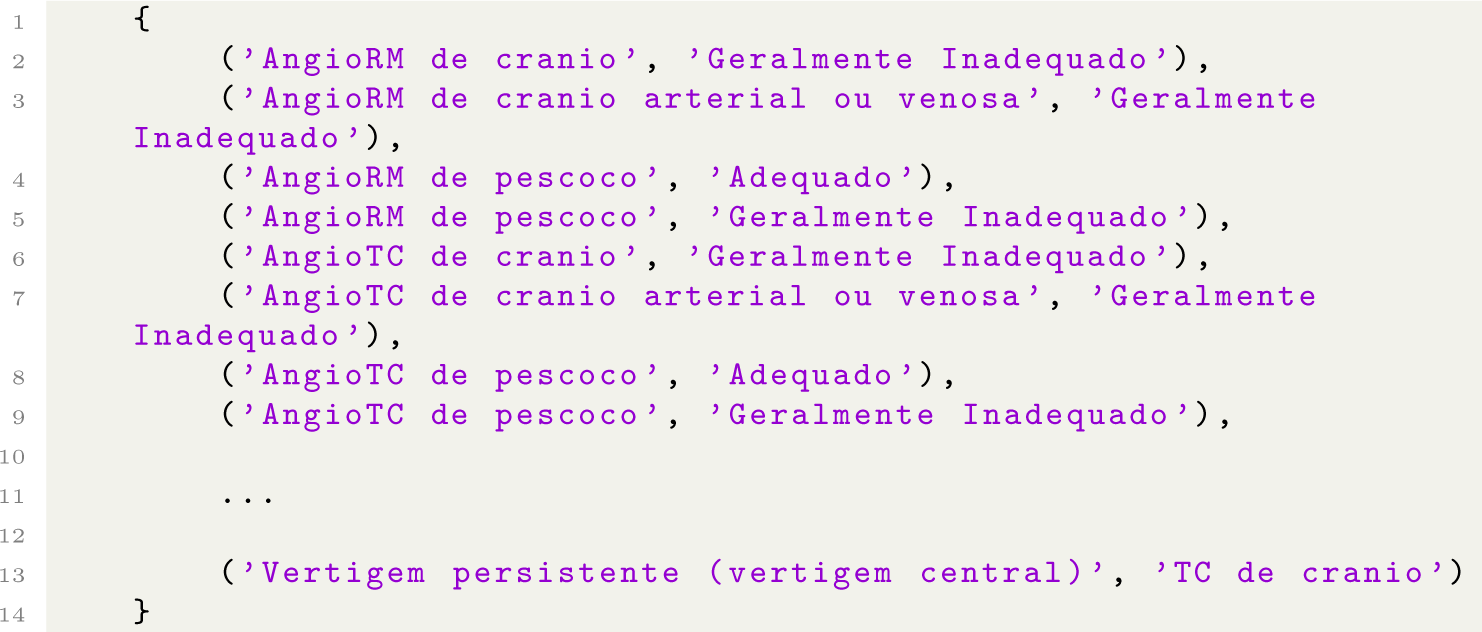
Output obtained for Experiment 3.

## 7 Conclusion

In this study we have explored an integrative approach bridging aspects of classical decision tables and modern machine learning, yielding simpler and sound approach to knowledge modeling for clinical decision support systems. We provide straightfoward mathematical results and computational algorithms, enforcing that valuable theoretical and practical findings can be obtained by intersectioning well-maturated artificial intelligence research and current machine learning formalisms. Through a python implementation, a real case scenario is used to ilustrate the application of our results in a knowledge modeling problem for medical radiology immaging exam prescription based on guidelines data. Future work on benchmarking the proposed algorithms embodied in a fully operational clinical decision support system could extend our findings towards daily used systems.

## Data Availability

All data produced in the present study are available upon reasonable request to the authors.

https://acsearch.acr.org/list

---

**Figure 1:**
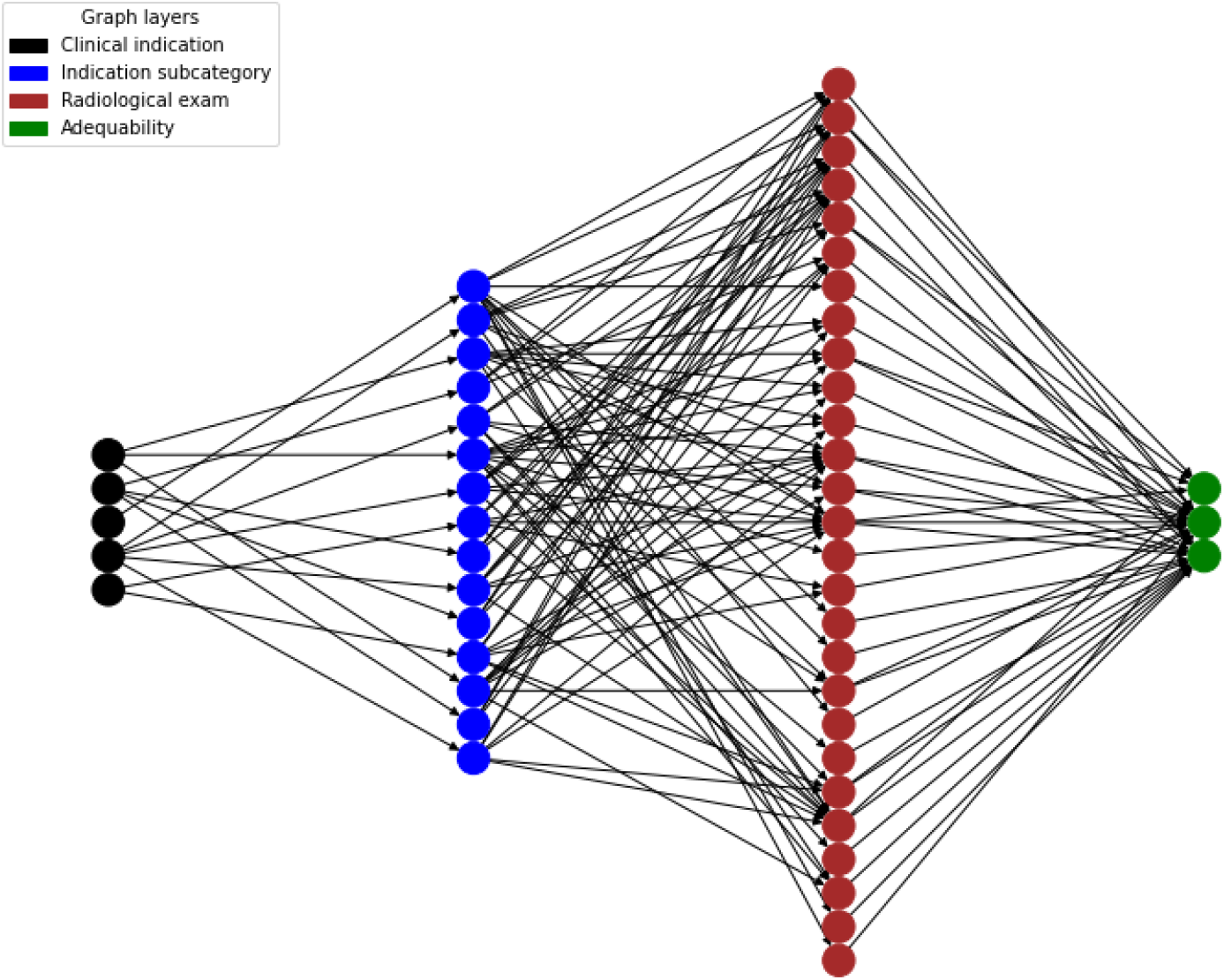
Graph representation for our experimental decision table, obtained using networkx package for the output of *from paths to edges*.

## Notes

### Competing Interest Statement

The authors have declared no competing interest.

### Funding Statement

This study was funded by Sociedade Beneficente Israelita Brasileira Albert Einstein.

